# Closing the Paediatric Gap: Adult-Trained AI Generalises Robustly to Paediatric Coeliac Disease Diagnosis

**DOI:** 10.64898/2026.06.04.26354889

**Authors:** Florian Jaeckle, Peter M. Gillett, Kathryn J. Kirkwood, Shonali Natu, James Y. H. Chan, Adrian C. Bateman, Mark J. Arends, Elizabeth J. Soilleux

**Affiliations:** Department of Pathology, University of Cambridge, Cambridge, UK; Lyzeum Ltd, Cambridge, UK; Department of Child Life and Health, University of Edinburgh, Royal Hospital for Children and Young People, UK; Department of Pathology, Western General Hospital, Edinburgh, UK; University Hospital of North Tees, North Tees and Hartlepool NHS Foundation Trust; Cambridge University Hospitals NHS Foundation Trust, Cambridge, UK; University Hospital Southampton NHS Foundation Trust, Southampton, UK; Edinburgh Pathology & Centre for Comparative Pathology, Institute of Genetics & Cancer, University of Edinburgh, Edinburgh, UK

## Abstract

**Background:** Coeliac disease (CD) diagnosis on duodenal biopsies is limited by interobserver variability. We have previously demonstrated pathologist-level performance with our artificial intelligence (AI) model for the histopathological diagnosis of adult CD, but not in paediatric practice. As paediatric CD screening programmes expand internationally, accurate and scalable diagnostic tools are needed. We investigated whether an AI model trained exclusively on adult whole-slide images (WSIs) can generalise to paediatric CD diagnosis across independent centres.

**Methods:** A training and validation dataset of 9,958 WSIs from 8,421 adult patients (961 CD) from five centres was used to develop an ensemble of multiple-instance learning models using features from a foundation model. Testing was performed on 708 consecutive paediatric patients (86 CD) from two centres (Edinburgh and Southampton) not included in training. Model calibration was assessed, and probability outputs were grouped into clinically interpretable categories.

**Findings:** In adult cross-validation, the AI model achieved an area under the receiver operating characteristic curve (AUC) of 98.7%, sensitivity of 84.9%, specificity of 99.0%, and negative predictive value (NPV) of 98.1%. On testing (paediatric) datasets, performance remained high (AUC 98.8%, sensitivity 80.2%, specificity 98.4%, NPV 97.3%). Restricting analysis to predictions outside the intermediate-probability range (predicted CD probability <10% or ≥65%; 85.3% of cases) improved sensitivity to 100% and specificity to 98.7%. No misclassifications were observed among high-confidence predictions (<2% or ≥85%; 66.0% of cases). The expected calibration error was 0.03. Performance improved significantly when biopsies from both duodenal sites (bulb [D1] and descending [D2/3]) were considered.

**Interpretation:** Our AI model, trained on adult biopsies, generalises to paediatric CD diagnosis across centres and scanner platforms. Well-calibrated probability outputs provide clinically interpretable measures of diagnostic confidence and could support safe identification of CD-negative biopsies within defined thresholds. These findings demonstrate the feasibility of applying adult-derived AI models in paediatric populations and reinforce the importance of multi-site (D1 & D2) biopsy sampling.

## Introduction

Coeliac disease (CD) is a common autoimmune disorder triggered by gluten ingestion affecting approximately 1% of the global population, yet the majority of cases remain undiagnosed [1,2]. Symptoms range from classical features of malabsorption, including chronic diarrhoea, weight loss, anaemia, and fatigue, to extra-intestinal manifestations such as dermatitis herpetiformis and infertility [3]. If left untreated, CD carries a significantly elevated risk of serious long-term complications, including lymphoma and adenocarcinoma [3,4]. Beyond these physical manifestations, coeliac disease particularly in children and adolescents is increasingly recognised to carry substantial psychosocial burden, including anxiety, food-related hypervigilance, social restriction, and impaired quality of life associated with maintaining a strict gluten-free diet [5,6], highlighting the importance of an accurate diagnosis.

The gold standard for diagnosis, as recommended by NICE, is histological examination of duodenal biopsies [7], however, this process is subject to significant interobserver variability as acknowledged by the recent European Society guidelines [8]. The mean probability of two pathologists agreeing on a CD diagnosis from digitised whole-slide images has been shown to be as low as 0.73, with a Cohen’s kappa of 0.59 in the absence of clinical or laboratory data, improving only modestly to 0.80 when serological information is included [9–12]. This diagnostic inconsistency has significant implications for patient care and underscores the need for more objective and reproducible methods of biopsy assessment.

Recent advances in artificial intelligence have shown considerable promise in addressing this challenge. In our previous multi-centre study, we demonstrated that our AI models achieve expert pathologists-level accuracy in detecting CD from duodenal biopsies in adult patients [13]. Although multiple studies have explored AI-based approaches to CD diagnosis [13–17] the paediatric domain remains underexplored. Syed et al. [18] attempted to address this gap by developing a convolutional neural network to classify CD, environmental enteropathy (EE), and normal duodenal biopsies in children; however, the study has a critical methodological limitation: EE images were sourced from hospitals in Pakistan and Zambia, while CD and normal images came from a different institution in the United States, and the two groups were acquired using different imaging methods. As the disease label and image source are therefore entirely confounded, it is impossible to determine whether the proposed model learned to identify pathological features or simply to recognise the origin of the images. To our knowledge, no study to date has demonstrated pathologist-level AI performance specifically in paediatric CD biopsies. Furthermore, some authors have suggested that paediatric and adult coeliac disease may differ in their biological and histopathological characteristics, with reported differences in gene expression [19] as well as in clinical presentation and mucosal pathology across age groups [20–23].

There is an increasing gap between the amount of AI research in paediatric coeliac disease compared to adult populations. Mass screening strategies for paediatric CD have been reported across 17 countries [24], most notably in Italy, which has become the first country to mandate nationwide screening for coeliac disease in children. A pilot programme has screened over 5,000 children aged 2–3, 6–7, and 10–11 years across four regions [25], using serological testing for coeliac-specific autoantibodies, including anti-tissue transglutaminase IgA with concurrent assessment of total IgA. Children with positive screening results are referred for specialist evaluation, where diagnosis is confirmed according to established paediatric guidelines through repeat serology and, in cases not meeting serology-based diagnostic criteria (e.g. anti-tissue transglutaminase IgA levels below ten times the upper limit of normal [26]), duodenal biopsy sampling from both the bulb (D1) and descending duodenum (D2), as recommend by the 2020 European Society Paediatric Gastroenterology, Hepatology and Nutrition (ESPGHAN) Guidelines for Diagnosing Coeliac Disease [26]. This multi-site approach is particularly important given the recognised existence of ultra-short coeliac disease, in which villous atrophy is confined to the duodenal bulb and may be missed by descending duodenal sampling alone [27,28]. As programmes such as these expand internationally, the number of children undergoing confirmatory diagnostic assessment, including histological evaluation, will increase, driving demand for accurate, scalable tools to support CD diagnosis on paediatric biopsies.

Despite this growing clinical need, children remain profoundly underrepresented in medical AI development. A cross-sectional analysis of 876 FDA-authorised AI and machine learning-enabled medical devices found that only 17% were explicitly labelled for use in children, and of those, only one in five had reported development using paediatric datasets [29]. A separate review of AI medical devices in radiology found that only 4 of 213 devices were clearly labelled for paediatric use [30]. This regulatory gap exposes children to the risk of off-label use of tools validated only on adult populations, a concern that extends well beyond radiology. Children are not simply small adults: physiological, anatomical, and histological differences mean that AI tools trained exclusively on adult data may fail or perform sub-optimally when applied to paediatric cases, a risk that remains poorly characterised for the vast majority of deployed tools.

Against this backdrop, we investigate whether an AI model trained exclusively on adult whole slide images (WSIs) can generalise to paediatric CD diagnosis across unseen clinical centres, and whether key histomorphological features of CD are sufficiently conserved across age groups to permit this transfer of learning.

## Methods

### Dataset and study cohorts

#### Training and validation dataset

The training and validation dataset comprised haematoxylin and eosin-stained duodenal biopsy whole-slide images from 8421 adult patients (961 coeliac disease, 6262 normal and 1198 with another diagnosis; ≥18 years old) collected from five hospitals, scanned at 40x magnification (∼0.25μm per pixel) using five different scanners from four manufacturers. Most contributing centres provided a series of consecutive biopsies, after which datasets were enriched for coeliac disease cases and, in some instances, for other non-coeliac pathological abnormalities to ensure adequate representation of diagnostically relevant classes (Table 1). Supplementary Materials S1 contains a detailed breakdown of the non-coeliac, but pathological, diagnoses.

**Table 1.** Training and testing datasets. Training data were collected from five UK centres and includes adult patients only. Test data were collected from two different UK centres and include paediatric cases only. Scanner model and patient age and sex are shown for both datasets. “Other” diagnoses comprise 14 different conditions; see Supplementary Materials S1 for a full breakdown of these diagnoses.

|  |  | Training Data |  |  | Testing Data |  |  |
| --- | --- | --- | --- | --- | --- | --- | --- |
|  |  | Coeliac<br>(n =<br>961) | Normal<br>(n =<br>6262) | 'Other'<br>(n =<br>1198) | Coeliac<br>(n =<br>86) | Normal<br>(n =<br>479) | 'Other'<br>(n =<br>143) |
| Source | Addenbrookes | 201 | 3228 | 777 | - | - | - |
|  | North Tees | 16 | 1517 | 172 | - | - | - |
|  | Heartlands | 633 | 563 | 0 | - | - | - |
|  | Glasgow | 70 | 538 | 34 | - | - | - |
|  | Warwick | 41 | 416 | 215 | - | - | - |
|  | Edinburgh | - | - | - | 55 | 159 | 23 |
|  | Southampton | - | - | - | 31 | 320 | 120 |
| Scanner | Leica Aperio AT2 | 273 | 1008 | 181 | - | - | - |
|  | Hamamatsu Nanozoomer XR | 194 | 187 | 0 | - | - | - |
|  | Hamamatsu Photonics K.K. | 16 | 1517 | 172 | - | - | - |
|  | Roche Ventana iScan HT | 245 | 189 | 0 | - | - | - |
|  | Philips IntelliSite Ultra Fast Scanner | 233 | 3361 | 845 | 55 | 159 | 23 |
|  | 3DHISTECH Kft. Panoramic 1000 | - | - | - | 31 | 320 | 120 |
| Sex | Female | 632 | 3608 | 557 | 57 | 229 | 60 |
|  | Male | 329 | 2651 | 640 | 29 | 248 | 80 |
| Age | median | 44 | 64 | 66 | 12 | 13 | 13 |
|  | 0-5 | - | - | - | 10 | 52 | 15 |
|  | 6-10 | - | - | - | 26 | 106 | 34 |
|  | 11 - 17 | - | - | - | 50 | 313 | 94 |
|  | 18 - 29 | 195 | 348 | 47 | - | - | - |
|  | 30 - 39 | 191 | 437 | 89 | - | - | - |
|  | 40 - 49 | 184 | 727 | 119 | - | - | - |
|  | 50 - 59 | 116 | 1017 | 202 | - | - | - |
|  | 60 - 69 | 146 | 1363 | 255 | - | - | - |
|  | 70 - 79 | 82 | 1524 | 315 | - | - | - |
|  | 80 - 89 | 46 | 797 | 157 | - | - | - |
|  | 90 - 99 | 1 | 49 | 14 | - | - | - |

#### Testing datasets

Two independent external testing cohorts were used: one from Southampton General Hospital in England and one from Western General Hospital in Edinburgh, Scotland, providing geographic and technical heterogeneity. The Edinburgh cohort was digitised using the same scanner platform as one of the training centres, whereas the Southampton cohort was scanned using a scanner from a different manufacturer, enabling assessment of cross-scanner generalisability. The combined testing datasets contain biopsies from 708 unique paediatric patients (86 coeliac disease, 479 normal, 143 other diagnosis). The mean patient age was 13 years (range 1–17 years), and 49% of patients were female (Table 1). For the entire Edinburgh cohort and the majority of the Southampton cohort, biopsies performed for suspected coeliac disease included samples from both the first (D1) and second (D2) portions of the duodenum, in line with routine clinical practice. The test dataset comprised consecutive cases and is representative of routine clinical practice. Patients undergoing repeat biopsy on a gluten-free diet were excluded from analysis.

#### Reference standard and diagnostic classification

For the training dataset, reference diagnoses were derived from the original histopathology report. Cases in which the report indicated a high-confidence diagnosis were labelled directly. Where histopathological findings were equivocal or insufficient for confident classification, additional clinical information was reviewed, including coeliac serology and independent assessment by a second pathologist, to establish a reference diagnosis.

For the testing datasets, an equivalent approach was used for the Southampton cohort. For the Edinburgh cohort, reference diagnoses were established through a multidisciplinary clinical consensus process involving paediatric gastroenterologists and pathologists. This process integrated histopathological findings, serological results, and relevant clinical information, including patient history and endoscopic findings, to determine the final clinical diagnosis. All reference diagnoses were assigned independently of AI model outputs.

### Image pre-processing and feature extraction

Whole-slide images underwent automated tissue detection and artefact removal using the method described by Schreiber *et al*. [31]. Tissue regions were divided into non-overlapping 256 × 256-pixel patches at 10x magnification. Stain variability was reduced using Macenko stain normalisation [31]. Patch-level feature embeddings were extracted using the hibou-b foundation model [32], which was used as a fixed feature extractor.

### Model development and training

We trained four independent binary classification models to distinguish coeliac disease-positive from coeliac disease-negative biopsies. AI model development employed leave-one-source-out cross-validation, whereby models were trained on data from all but one contributing centre and evaluated on the held-out source, enabling assessment of robustness to institutional variation. All experiments were repeated using three random seeds to assess training stability. Models were implemented using the CLAM, ABMIL, and TransMIL multiple-instance learning architectures [33–35]. During training, data sampling strategies were applied to address imbalances in diagnostic categories and contributing centres; full details are provided in Supplementary Material S2.

### Model validation and calibration

For each trained classifier, predicted CD probabilities were recalibrated using validation data to improve how well predicted risks matched observed outcomes. To ensure consistency at the patient level, only the scan with the highest abnormal score per case was used for calibration (reflecting clinical practice, where the most abnormal site is typically reported). The recalibration approach also accounted for differences between the prevalence of disease in the training data and that expected in real-world clinical settings.

### Model inference

At inference time, predictions were generated using an ensemble of all trained classifiers. Calibrated probabilities from each individual model were averaged to produce a final mean probability for each case. Ensembling reduced the frequency of extreme predicted probabilities and improved prediction stability. The confidence thresholds below were used to create a final diagnosis.

### Diagnostic categories

To support clinical interpretation, predicted coeliac disease probabilities (p) were grouped into five categories using predefined confidence thresholds: very unlikely coeliac disease (p<2%), unlikely coeliac disease (2%≤p<10%), intermediate probability (10%≤p<65%), likely coeliac disease (in the appropriate clinical and serological context) (65%≤p<85%), and very likely coeliac disease (p≥85%).

Thresholds were selected pragmatically to favour sensitivity over specificity, reflecting the greater clinical concern associated with false-negative predictions and the intended use of the AI model as an adjunct to serological and histopathological assessment in a condition that remains underdiagnosed. Lower thresholds were aligned with typical pre-test probability in investigated populations (approximately 5–10% [36]) such that predictions below this range do not increase suspicion beyond baseline risk. Higher thresholds represent increasing levels of diagnostic confidence, with “likely” and “very likely” categories intended to support clinical decision-making in the appropriate clinical and serological context.

### Performance metrics

AI model performance was assessed using accuracy, sensitivity, specificity, positive and negative predictive value (PPV and NPV), and the area under the receiver operating characteristic curve (AUC) defined according to standard epidemiological conventions. Differences in AUC between AI models were compared using the DeLong test, with a two-sided p-value <0.05 considered statistically significant [37]. AI model calibration was evaluated using expected calibration error (ECE), which quantifies the agreement between predicted probabilities and observed outcome frequencies.

## Results

### Adult cross-validation

We trained four independent AI models on a dataset of duodenal biopsies from 8421 adult patients (≥18 years) and assessed performance using leave-one-source-out cross-validation. For patients with multiple whole-slide images (WSIs), for example, biopsies obtained from both the duodenal bulb (D1) and descending duodenum (D2/3), or multiple sections prepared from the same biopsy, each WSI was analysed independently to generate a predicted probability (risk estimate) of coeliac disease. At the patient level, the maximum predicted coeliac disease probability across all WSIs was used to give the overall diagnostic classification, reflecting a clinically conservative approach in which a single highly suspicious biopsy is sufficient to support the diagnosis.

Leave-one-source-out cross-validation on our dataset of 8421 adult patients showed that our AI model achieved an overall validation accuracy of 97.4%, sensitivity of 84.9%, and specificity of 99.0% across all cases. Restricting analysis to high-confidence predictions (predicted CD probability p<2% or p≥85%; 83.1% of cases) improved performance to 99.3% accuracy, 97.7% sensitivity, and 99.5% specificity (Table S2 in Supplementary Materials S2).

The AI model achieved near identical performance for male and female patients (AUC 98.6% vs 98.7%, respectively) and similarly performance across all adult age groups (AUC 96.7%-100%) (Table S2).

The expected calibration error (ECE) was 0.005, indicating that, on average, predicted probabilities for coeliac disease differed from observed outcomes by about 0.5 percentage points. In practical terms, a predicted risk of 60% would typically correspond to an observed disease frequency of around 59.5–60.5%, although variation around this is expected. This reflects close agreement between predicted likelihood of disease and actual disease frequency, suggesting that the AI model’s probability estimates are reliable for clinical interpretation. However, this estimate may be optimistic, as model calibration (i.e., agreement between predicted probabilities and observed outcomes) was evaluated on the same validation dataset used to fit the model.

### Paediatric testing

We evaluated the AI model, trained exclusively on adult patients, on an independent paediatric dataset comprising duodenal biopsies from 708 unique patients (aged 1–17 years; mean age 13; 49% female) from two hospitals not included in the training data, providing an independent test of generalisability. The test dataset comprised consecutive cases and is representative of routine clinical practice.

On this dataset the AI model achieved an accuracy of 96.2%, sensitivity of 80.2%, and specificity of 98.4%. Restricting analysis to predictions outside the intermediate-probability range (predicted CD probability <10% or ≥65%; retaining 85.3% of cases), performance improved to 98.8% accuracy, 100% sensitivity, and 98.7% specificity. Among high-confidence predictions (defined as “very unlikely” or “very likely” coeliac disease; p<2% or p≥85%; 66% of cases retained), no misclassifications were observed (Table 2).

**Table 2.** Test set results on the paediatric dataset. Exclude intermediate-probability predictions leaves cases with a predicted CD probability of <0.10% or ≥0.65%. High-confidence predictions include CD probabilities of <2% or ≥85%. All subgroup analysis is done on all cases including ones within the intermediate-probability range. PPV = positive predictive value. NPV = negative predictive value. AUC = area under the ROC curve.

|  |  | Accuracy | Sensitivity | Specificity | PPV | NPV | AUC |
| --- | --- | --- | --- | --- | --- | --- | --- |
|  | Overall (n=708) | 96.2 | 80.2 | 98.4 | 87.3 | 97.3 | 98.8 |
|  | Excluding intermediate-probability predictions (n=604, 85.3%) | 98.8 | 100 | 98.7 | 89.1 | 100 | 99.6 |
|  | High-confidence predictions (n=467, 66.0%) | 100 | 100 | 100 | 100 | 100 | 100 |
| Source | Edinburgh (n=237) | 94.1 | 81.8 | 97.8 | 91.8 | 94.7 | 98.4 |
|  | Southampton (n=471) | 97.2 | 77.4 | 98.6 | 80.0 | 98.4 | 99.0 |
| Sex | Male (n=357) | 96.9 | 79.3 | 98.5 | 82.1 | 98.2 | 98.6 |
|  | Female (n=349) | 95.4 | 80.7 | 98.3 | 90.2 | 96.3 | 98.9 |
| Age | 1-5 (n=77) | 98.7 | 90.0 | 100.0 | 100.0 | 98.5 | 99.4 |
|  | 6-10 (n=166) | 92.8 | 76.9 | 95.7 | 76.9 | 95.7 | 97.7 |
|  | 11-17 (n=457) | 96.9 | 80.0 | 99.0 | 90.9 | 97.6 | 99.0 |

Performance was consistent across centres, sexes, and age groups (Table 2), indicating robust generalisability across clinically relevant subgroups.

Calibration remained strong in the paediatric cohort, with an expected calibration error of 0.03, indicating that, on average, predicted probabilities for coeliac disease differed from observed outcomes by approximately 3 percentage points. In practical terms, this means that if the AI model predicts a 70% probability of disease, the true proportion of patients with disease would generally be within 3% of this value. This reflects close agreement between predicted likelihood and observed disease frequency (Figure 2).

**Figure 1.**
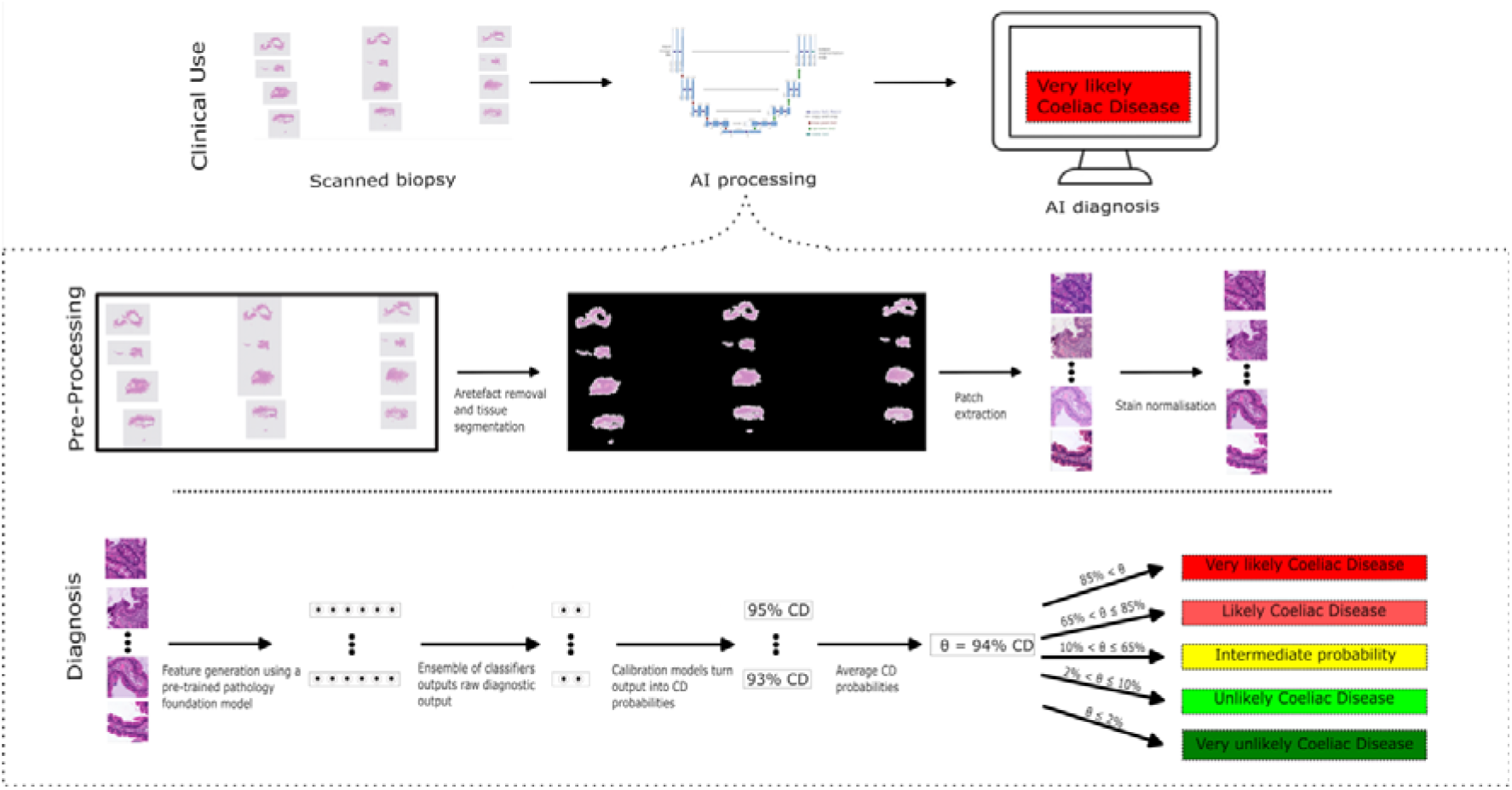
Model pipeline. The pipeline consists of the following steps: (1) digitised whole-slide images of duodenal biopsies are received; (2) artefacts are removed and tissue regions are identified; (3) tissue regions are divided into small image patches; (4) stain normalisation is applied; (5) patch-level features are extracted using a pre-trained pathology model; (6) an ensemble of classifiers generates slide-level predictions from patch-level features; (7) model outputs are calibrated to produce reliable probabilities of coeliac disease; (8) calibrated probabilities are averaged to generate a final patient-level probability; and (9) this probability is mapped to clinically interpretable diagnostic categories (very unlikely, unlikely, intermediate-probability, likely, and very likely coeliac disease). The final output is presented as a probability-informed diagnosis to support clinical decision-making.

**Figure 2.**
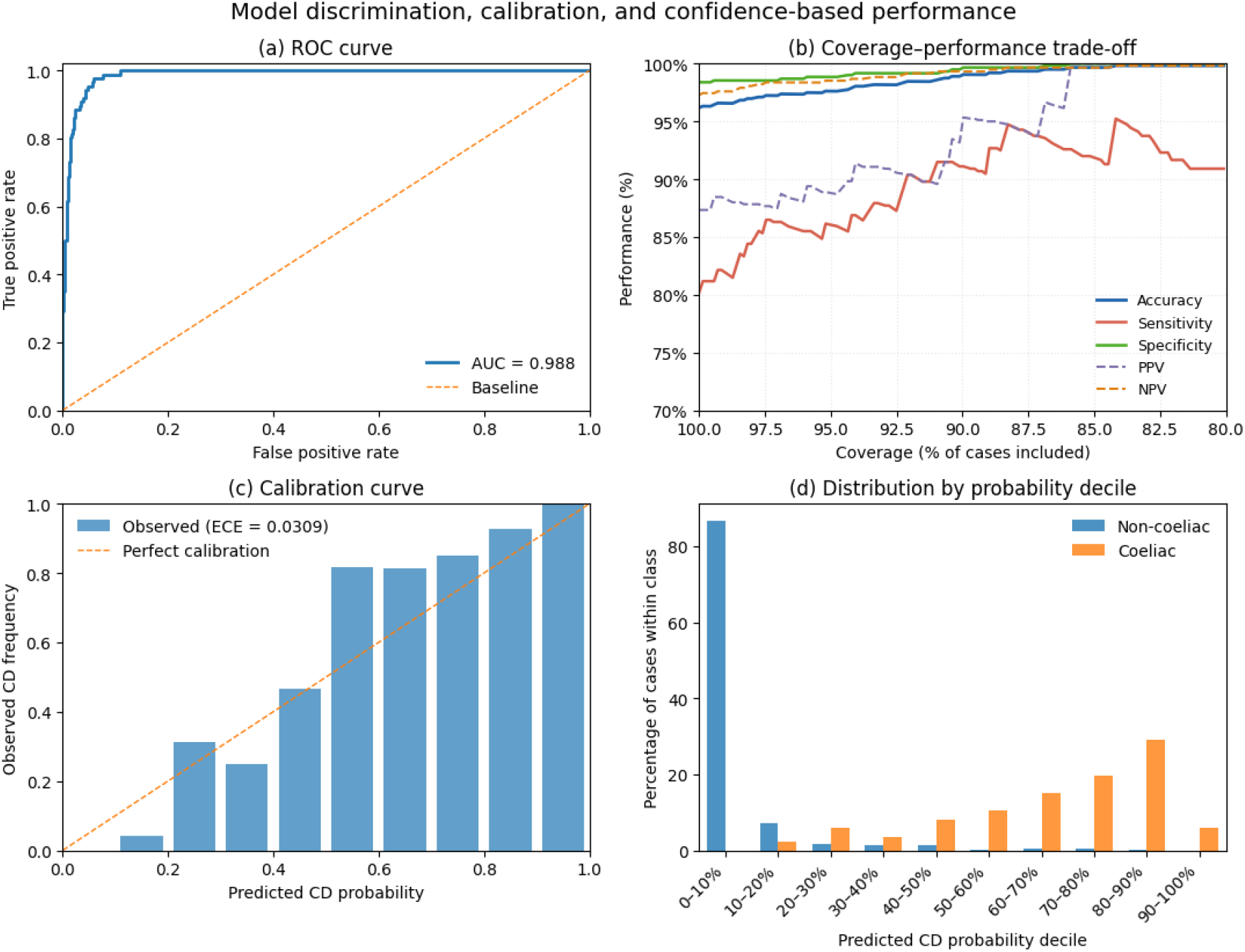
Model discrimination, calibration, and confidence-based performance. (a) Receiver operating characteristic (ROC) curve demonstrating excellent discrimination of coeliac disease (area under the curve (AUC) = 0.988). (b) Coverage–performance trade-off using symmetric probability thresholds (including cases with predicted probability <t or >1™t). Restricting analysis to the most confident predictions improved performance, with accuracy approaching ∼99% at approximately 90% coverage. Sensitivity and specificity remained high across the range of coverage. (c) Calibration curve showing good agreement between predicted and observed probabilities (expected calibration error [ECE] = 0.0309), indicating that predicted probabilities are well calibrated. (d) Distribution of predicted probabilities by class, demonstrating separation between coeliac and non-coeliac cases, with most non-coeliac cases assigned low probabilities and coeliac cases concentrated at higher probabilities.

### Contribution of duodenal biopsy site (D1 vs D2)

We next assessed the contribution of biopsy location using paired samples from the first (D1) and second (D2) parts of the duodenum (Figure 3) in a subset of the test dataset (226 biopsies from 113 patients in the Edinburgh cohort with suspected coeliac disease, of whom 55 (48.7%) were diagnosed with coeliac disease). Diagnostic performance was lower when using either site in isolation (AUC of 87.1% for D2 and 91.8% for D1) than when combining information from both biopsies. When the predicted probabilities of coeliac disease from D1 and D2 were averaged to derive a patient-level prediction, the AUC was 96.6%; when the maximum predicted probability across the two sites was used (as in the primary analysis), the AUC was 95.7%.

**Figure 3.**
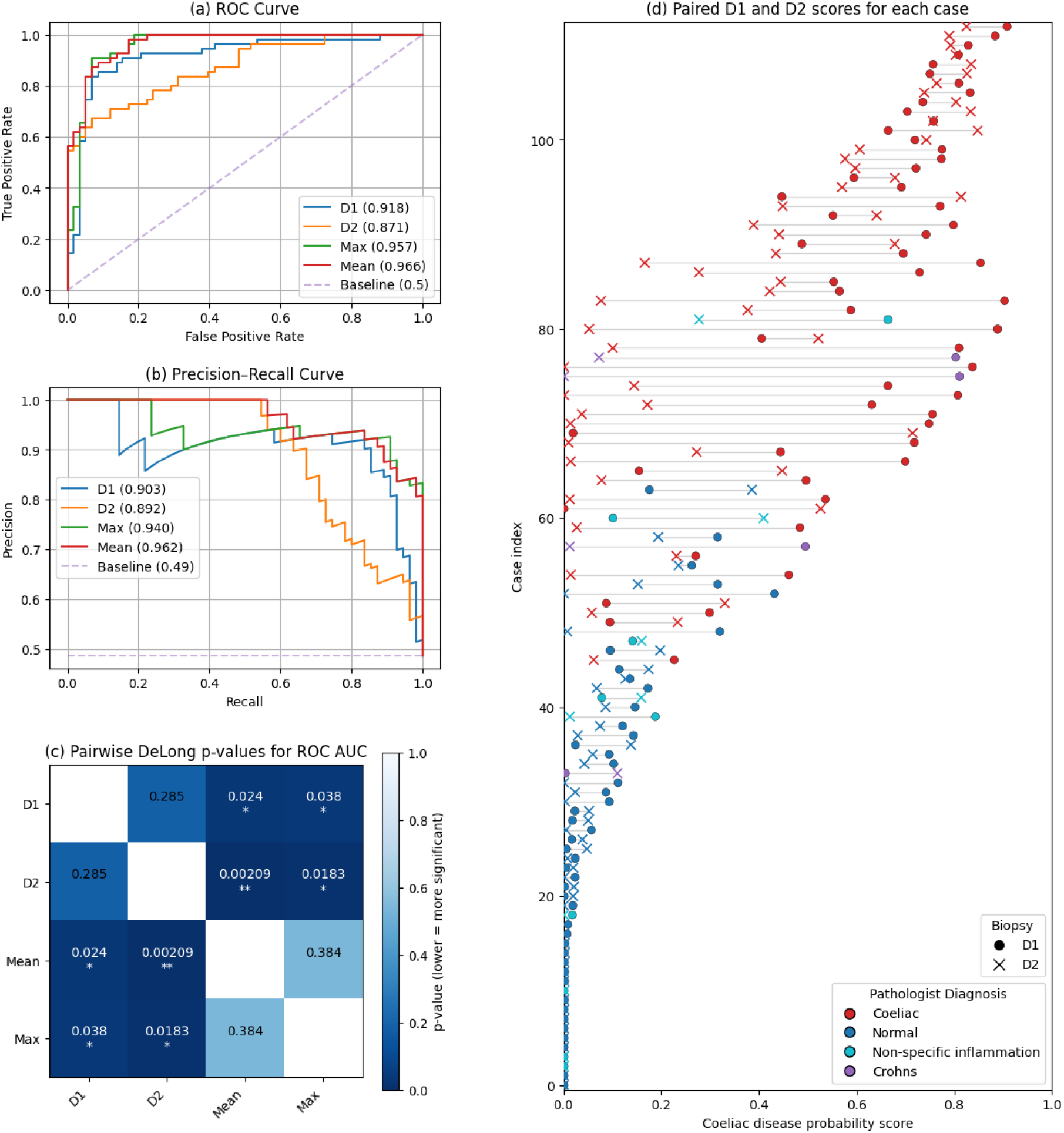
Performance of the AI model on paired D1 and D2 duodenal biopsies. (a) Receiver operating characteristic (ROC) curves for individual biopsy scores (D1 and D2) and combined scores (maximum and mean across biopsies). The mean and maximum scores showed higher discrimination than either biopsy alone. (b) Precision–recall curves for the same models, with the baseline indicating disease prevalence. Performance trends were consistent with ROC analysis. (c) Pairwise comparisons of ROC area under the curve (AUC) using DeLong’s test. Lower p values (darker shading) indicate stronger evidence of differences between models. Combined scores were significantly better than D2 and modestly better than D1, with no significant difference between mean and maximum aggregation. (d) Paired comparison of per-patient scores from the two biopsies. Each line represents an individual, linking D1 (circle) and D2 (cross) scores, coloured by histopathological diagnosis. Variation between biopsies within the same patient is evident, and combined scoring reduces this variability.

These differences were statistically significant (DeLong test). For D2, performance was lower than both the mean-based (p<0.003) and max-based (p<0.02) combined approaches; similarly, for D1, performance was lower than the mean-based (p<0.03) and max-based (p<0.04) approaches. This highlights the added diagnostic value of multi-site sampling. Analysis of paired predictions further demonstrated substantial intra-patient variability in predicted coeliac disease probability between D1 and D2 samples (Figure 3D).

## Discussion

In this multi-centre study, we found that an AI model trained exclusively on adult duodenal biopsy whole-slide images can generalise effectively to paediatric coeliac disease diagnosis, achieving high accuracy on independent external datasets. Importantly, these datasets incorporated both inter-institutional variation and differences in scanning platforms, providing a test of real-world generalisability. Despite being trained solely on adult data, the AI model maintained strong performance in children, a population that is frequently underrepresented in medical artificial intelligence development. This finding is particularly relevant given the practical constraints of paediatric data collection, where limited biopsy availability often restricts AI model development. Our results suggest that adult-derived datasets may be sufficient for training robust AI models in coeliac disease, allowing scarce paediatric biopsies to be reserved for independent evaluation and validation rather than training.

Beyond overall diagnostic accuracy, our study emphasises the importance of reliable probability estimates for clinical deployment. In routine practice, diagnostic decisions are rarely binary and instead depend on the degree of certainty associated with a given finding. AI models have been criticised for producing probability estimates that do not reflect true likelihood, for example, assigning very high confidence to incorrect predictions, which limits their clinical utility. In contrast, our AI model’s predicted probabilities closely matched the observed frequency of disease, meaning that cases assigned a high probability of coeliac disease were very likely to be truly positive, and vice versa. Notably, predictions classified by the AI model as “very likely coeliac disease” or “very unlikely coeliac disease” were all correctly classified within this dataset and accounted for two thirds of cases in the paediatric cohort. This ability of an AI model to provide reliable and clinically interpretable measures of diagnostic confidence is essential for integration into clinical workflows.

Our findings also reinforce the continued importance of established biopsy protocols. Current guidelines from the European Society Paediatric Gastroenterology, Hepatology and Nutrition [26] and the North American Society For Pediatric Gastroenterology, Hepatology & Nutrition [38] recommend sampling from both the duodenal bulb (D1) and descending duodenum (D2) to maximise diagnostic yield. This is particularly relevant given the recognised existence of ultra-short coeliac disease, in which villous atrophy is confined to the duodenal bulb, and which may be missed by descending sampling alone [27,28]. We demonstrate that this principle remains highly relevant in the context of AI-assisted assessment. AI model performance was significantly improved when both D1 and D2 biopsies were available compared with either site alone, with statistically significant gains in discriminative ability. Furthermore, we observed substantial variability in predicted disease probability between paired D1 and D2 samples within the same patient, underscoring the patchy nature of coeliac disease and the risk of sampling error. These findings suggest that adherence to current biopsy guidelines will be critical for ensuring optimal performance of AI tools in clinical practice and provide independent empirical support for existing recommendations to sample both D1 and D2.

This study has several limitations. First, all data were derived from UK centres; although performance was consistent across multiple institutions and scanner types, generalisability to other populations and health-care settings remains uncertain. Second, the reference standard for coeliac disease is imperfect, with recognised interobserver variability in histopathological interpretation. Use of rigorous multidisciplinary consensus between gastroenterologists and pathologists was undertaken, aiming to improve diagnostic accuracy, but misclassification of a small proportion of biopsies still cannot completely be excluded.

Taken together, these findings support the feasibility of deploying AI-based approaches for coeliac disease assessment across age groups and clinical settings. The results also highlight important considerations for safe implementation, including the need for well-calibrated probability outputs (i.e., predicted coeliac disease probabilities that closely reflect the true likelihood of disease) and adequate biopsy sampling (including sampling from both the duodenal bulb (D1) and descending duodenum (D2), in line with current guidelines [26,38]). High accuracy among high-confidence predictions suggests that a threshold-based approach could enable AI-assisted identification of biopsies that are very unlikely to represent coeliac disease, potentially supporting safe workload reduction in pathology through automated triaging or fully automated diagnostic reporting of such cases.

The clinical importance of accurate and timely diagnosis extends beyond histopathology alone. Increasing evidence demonstrates substantial psychosocial burden among children and adolescents with coeliac disease, including anxiety, social restriction, impaired quality of life, and hypervigilance surrounding gluten exposure [5,6,39]. Prolonged undiagnosed disease is additionally associated with increased long-term morbidity [40,41]. Ongoing debate regarding population-based screening programmes is likely to further increase the number of children undergoing confirmatory endoscopic biopsy, driving demand for scalable and reproducible histopathological assessment.

Future work should include prospective evaluation of our AI model in routine diagnostic workflows, direct comparison with pathologist performance, and validation in more diverse international populations. Dedicated investigation of clinically important subgroups, including children with type 1 diabetes, selective IgA deficiency, and screen-detected asymptomatic cases, would help establish whether performance is maintained across the full spectrum of paediatric coeliac disease presentations.

## Conclusion

In this study, we showed that an AI model trained exclusively on adult duodenal biopsy whole-slide images can generalise effectively to paediatric coeliac disease diagnosis across independent clinical centres and scanning platforms. Despite well-documented concerns about the generalisation of adult-trained AI devices to paediatric populations, our findings suggest that key histomorphological features of coeliac disease appear sufficiently conserved to enable robust cross-age generalisation. This has important practical implications, particularly in the context of limited paediatric data availability, and supports a strategy in which paediatric datasets are prioritised for independent validation rather than model training.

Crucially, our AI model provides well-calibrated probability estimates, allowing uncertainty to be communicated in a clinically meaningful way. High-confidence predictions achieved perfect accuracy and accounted for the majority of cases, underscoring the potential for AI to support decision-making within existing diagnostic pathways. Together with the demonstrated importance of multi-site biopsy sampling, these findings suggest a pathway towards safe, scalable, and clinically integrated AI-assisted assessment of coeliac disease in children, with potential to support automated triaging and reporting of biopsies unlikely to represent coeliac disease.

## Data Availability

Data sharing statement: The data underlying this study are not publicly available.

## Acknowledgement

We would like to acknowledge Josephine Williams, Richard Walsh, Rosie Turnbull, Robert Richardson, Matthew Ellis, Singam Sivasankar, Michelle Watson, Avinash Thomas, Obinna Enwereji, Craig Marshall, and Vishad Patel for their support in collecting the data. We would further like to thank Graham Snudden for organisational support.

## Contributions

FJ, PG, MA, and ES conceived and designed the study. FJ developed the AI model, performed all experiments, and drafted the manuscript. KK, SN, JC, and AB oversaw data collection and clinical governance at their respective centres. PG, MA, and ES contributed to subsequent revisions and helped shape the final manuscript. All authors reviewed and approved the final version.

## Data sharing statement

The data underlying this study are not publicly available because the ethics approval and participant consent agreements do not permit data sharing.

