## Supplementary Materials for "Closing the Paediatric Gap: Adult-Trained AI Generalises Robustly to Paediatric Coeliac Disease Diagnosis"

### S1 Dataset

| Diagnosis | Training Dataset | Testing Dataset |
| --- | --- | --- |
| Normal | 6262 | 479 |
| Coeliac Disease | 961 | 86 |
| Chronic / non-specific inflammation | 428 | 103 |
| Adenoma | 182 | 0 |
| Isolated intestinal intraepithelial lymphocytes | 152 | 5 |
| Gastric Heterotopia | 113 | 0 |
| Gastric Metaplasia | 91 | 12 |
| Ulceration | 45 | 0 |
| Carcinoma | 35 | 0 |
| Peptic Duodenitis | 26 | 0 |
| Non-diagnosable | 21 | 8 |
| Lymphangiectasia | 19 | 0 |
| Isolated partial villous atrophy | 17 | 7 |
| Neuroendocrine Tumour | 9 | 0 |
| Transplant | 6 | 0 |
| Brunner Gland Hyperplasia | 5 | 0 |
| Giardia | 5 | 0 |
| Eosinophilic Duodenitis | 5 | 1 |
| Amyloidosis | 1 | 0 |
| Crohn’s Disease | 1 | 6 |
| Other Diagnosis | 20 | 0 |

Table S1. Detailed breakdown of diagnoses of the training and testing dataset.

#### S1.1 Adult Training Dataset

The training and validation dataset comprised duodenal biopsy whole-slide images from five UK centres, incorporating both consecutive and enriched cohorts to ensure representation of routine clinical practice alongside sufficient numbers of coeliac disease (CD) and other pathological cases.

At Addenbrooke’s Hospital, 4206 cases were included from two consecutive time periods: 2018–2019 (n=1206) and 2022–2024 (n=3000). These datasets were representative of routine clinical practice and were not enriched for any diagnostic category. The earlier cohort was scanned using the Leica Aperio AT2 scanner, and the later cohort using the Philips IntelliSite Ultra Fast Scanner.

From University Hospital of North Tees, 1705 cases were included, all of which were consecutive cases collected between 2022 and 2024 and scanned using a Hamamatsu NanoZoomer S210 scanner.

From Queen Elizabeth University Hospital, 642 consecutive cases representative of routine clinical practice were included, scanned using the Philips IntelliSite Ultra Fast Scanner.

From Warwick Hospital, 672 cases were included. Of these, 469 were consecutive cases representative of routine clinical practice and scanned using the Philips IntelliSite Ultra Fast Scanner. The remaining cases were selectively enriched to increase representation of coeliac disease and other non-coeliac pathological abnormalities and were scanned using either the Philips IntelliSite Ultra Fast Scanner or Leica Aperio AT2.

From Heartlands Hospital, 400 patients with either normal or coeliac disease diagnoses were included. These were scanned across three platforms (Hamamatsu NanoZoomer XR, Leica Aperio AT2, and Roche Ventana iScan HT), resulting in 1196 whole-slide images.

In total, the training and validation dataset comprised 6262 normal cases, 961 coeliac disease cases, and 1198 cases with other non-coeliac pathological diagnoses. A detailed breakdown of these non-coeliac diagnoses is provided in Table S1.

For the training dataset, reference diagnoses were derived from the original histopathology report. Cases in which the report indicated a high-confidence diagnosis were labelled directly. Where histopathological findings were equivocal or insufficient for confident classification, additional clinical information was reviewed, including coeliac serology and independent assessment by a second pathologist, to establish a reference diagnosis.

#### S1.2 Paediatric Testing Dataset

The paediatric test dataset comprised consecutive cases representative of routine clinical practice from two independent centres not included in model training.

From Western General Hospital, Edinburgh, 237 cases were included, scanned using the Philips IntelliSite Ultra Fast Scanner. From University Hospital Southampton, 471 cases were included, scanned using the 3DHISTECH Pannoramic 1000 scanner.

In total, the test dataset comprised 479 normal cases, 86 coeliac disease cases, and 143 cases with other non-coeliac pathological diagnoses. The distribution of these non-coeliac diagnoses differed from the training dataset, reflecting differences in patient age and underlying disease spectrum in paediatric populations. A detailed breakdown is provided in Table S1.

For data from the Southampton cohort, reference diagnoses were taken from the original histopathology report. For cases where the histopathological findings were equivocal, CD serology was also considered. For the Edinburgh cohort, reference diagnoses were established through a multidisciplinary clinical consensus process involving paediatric gastroenterologists and pathologists. This process integrated histopathological findings, serological results, and relevant clinical information, including patient history and endoscopic findings, to determine the final clinical diagnosis. All reference diagnoses were assigned independently of AI model outputs.

### S2 Methods

We performed four-fold leave-one-source-out cross-validation. Data from Glasgow and North Tees were combined due to their low number of coeliac disease (CD) cases. All experiments were repeated across three random seeds.

We trained multiple instance learning (MIL) models, including CLAM, ABMIL, and TransMIL (see Section S3.2). Models were trained for three epochs using the Adam optimizer with a constant learning rate of $2\times{10}^{-4}$ and weight decay of $1\times{10}^{-5}$.

Stain normalisation was applied using the Macenko method. To improve robustness, feature dropout was employed: for each case, between 20% and 70% of patch-level features were randomly sampled during training.

To address imbalance across data sources and class distributions, we assigned a sampling weight to each training example. For each source, a weight was computed proportional to the square root of its bottleneck class size (corresponding to the number of CD cases). This source-level weight was then evenly distributed across CD-positive and CD-negative samples within that source, ensuring equal contribution within each class. The resulting per-sample weights were normalised and used with a weighted sampling strategy during training to promote balanced learning across both sources and classes.

We trained a post-hoc calibration model to map raw model outputs to well-calibrated probabilities interpretable as absolute risk. Specifically, model scores were used as inputs to a LogisticRegression model, which learns a monotonic transformation from uncalibrated scores to predicted probabilities.

Because the development dataset was enriched for positive cases (observed prevalence ≈20%) relative to the expected real-world prevalence (10%), inverse probability weighting was applied during calibration. Each observation was assigned a weight proportional to the ratio of the target prevalence to the observed prevalence: positive cases were weighted by $0.1/0.2$, and negative cases by $0.9/0.8$. This reweighting ensures that the calibration model reflects the class distribution expected in clinical practice.

The calibrated model therefore yields probability estimates aligned with real-world event rates, rather than the artificially enriched distribution of the training data.

### S3 Validation performance

#### S3.1. Detailed Validation Performance by subcategory

The model achieved high validation performance on the adult dataset (n=8421), with an accuracy of 97.4% and AUC of 98.7%. Sensitivity was 84.9% and specificity 99.0%, indicating strong discrimination with high rule-out capability.

Restricting analysis to cases outside the intermediate-probability range (<10% or ≥65%; 93.9% of cases) improved performance (accuracy 98.8%, AUC 98.0), with further gains in high-confidence predictions (<0.02 or >0.85; 83.1% of cases), where sensitivity and specificity reached 97.7% and 99.5%, respectively (AUC 99.2%). Model performance increased with prediction confidence.

Performance was consistent across sites, sexes, and age groups. AUCs exceeded 98.6% across all centres, with some variation in sensitivity (79.6%–90.0%). Performance was similar between females and males, with modest differences in sensitivity (86.5% vs 81.8%). Across age groups, discrimination remained high (AUC 98–100%), although sensitivity was lower in older cohorts (e.g., 76.1% in ages 80–90). These findings indicate robust generalisation across diverse subgroups.

|  |  | Accuracy | Sensitivity | Specificity | PPV | NPV | AUC |
| --- | --- | --- | --- | --- | --- | --- | --- |
|  | Overall (n=8421) | 97.4 | 84.9 | 99.0 | 91.6 | 98.1 | 98.7 |
|  | Excluding intermediate-probability predictions (n=7911, 93.9%) | 98.8 | 94.5 | 99.3 | 93.8 | 99.4 | 98.0 |
|  | High-confidence predictions (n=6995, 83.1%) | 99.3 | 97.7 | 99.5 | 95.2 | 99.8 | 99.2 |
| Source | Addenbrookes (n=4206) | 98.0 | 79.6 | 99.0 | 79.2 | 99.0 | 98.6 |
|  | North-Tees (n=1705) | 98.7 | 87.5 | 98.8 | 41.2 | 99.9 | 99.5 |
|  | Warwick (n=672) | 97.9 | 90.0 | 98.4 | 78.3 | 99.4 | 99.0 |
|  | Heartlands (n=1196) | 92.4 | 85.6 | 100.0 | 100.0 | 86.1 | 99.6 |
|  | Glasgow (n=642) | 98.4 | 90.0 | 99.5 | 95.5 | 98.8 | 99.5 |
| Sex | Female (n=4799) | 97.5 | 86.5 | 99.2 | 94.1 | 98.0 | 98.7 |
|  | Male (n=3618) | 97.2 | 81.8 | 98.8 | 86.8 | 98.2 | 98.6 |
| Age | 18-30 (n=590) | 95.1 | 88.7 | 98.2 | 96.1 | 94.6 | 99.0 |
|  | 30-40 (n=717) | 94.1 | 83.8 | 97.9 | 93.6 | 94.3 | 98.5 |
|  | 40-50 (n=1030) | 95.6 | 81.0 | 98.8 | 93.7 | 96.0 | 96.7 |
|  | 50-60 (n=1335) | 98.4 | 88.8 | 99.3 | 92.8 | 98.9 | 98.9 |
|  | 60-70 (n=1764) | 98.3 | 87.7 | 99.3 | 91.4 | 98.9 | 99.1 |
|  | 70-80 (n=1921) | 98.1 | 81.7 | 98.9 | 76.1 | 99.2 | 99.0 |
|  | 80-90 (n=1000) | 98.4 | 76.1 | 99.5 | 87.5 | 98.9 | 98.1 |
|  | 90-100 (n=64) | 98.4 | 100.0 | 98.4 | 50.0 | 100.0 | 100.0 |

Table S2. Leave-one-source-out Cross-Validation Performance. Excluding intermediate probability range predictions meant keeping cases with a predicted CD probability <10% or ≥65%; High-confidence predictions include predicted CD probabilities of <2% or ≥85%. All subgroup analysis is done on all cases including ones within the intermediate probability.

#### S3.2. Validation performance – MIL comparison

|  | All cases | | Excluding intermediate-probability cases | | Very confident cases | |
| --- | --- | --- | --- | --- | --- | --- |
|  | percentage of total cases | AUC | percentage of total cases | AUC | percentage of total cases | AUC |
| CLAM | 100 | 98.7 | 93.9 | 99.0 | 83.1 | 99.2 |
| ABMIL | 100 | 99.1 | 94.5 | 99.3 | 84.4 | 99.6 |
| TransMIL | 100 | 98.8 | 93.3 | 99.1 | 84.9 | 99.2 |
| MIL-Ensemble | 100 | 99.1 | 94.0 | 99.4 | 85.0 | 99.6 |

Table S3. We compare performance across different MIL architectures. Performance is similar across all three MIL types.

We compared the performance of three multiple instance learning (MIL) architectures, CLAM, ABMIL, and TransMIL, and an ensemble combining all three approaches. To ensure a fair comparison, each MIL model was trained using three independent random seeds, and performance was averaged across runs. The ensemble was constructed by combining one trained model from each architecture and aggregating their predictions.

All models demonstrated high discrimination across evaluation settings. On the full dataset, AUCs ranged from 98.7% (CLAM, TransMIL) to 99.1% (ABMIL and ensemble). Performance improved when excluding intermediate-probability predictions, with AUCs increasing to 99.0–99.4%. Among very high-confidence cases, all models achieved excellent discrimination (AUC ≥99.2%), with ABMIL and the ensemble reaching the highest performance (99.6%).

Overall, differences between individual MIL architectures were modest, with consistently high performance across all methods. The ensemble provided a small but consistent improvement in discrimination, particularly in the high-confidence subset, suggesting complementary strengths across model types.
